# No evidence for allelic association between Covid-19 and ACE2 genetic variants by direct exome sequencing in 99 SARS-CoV-2 positive patients

**DOI:** 10.1101/2020.05.23.20111310

**Authors:** Antonio Novelli, Michela Biancolella, Paola Borgiani, Dario Cocciadiferro, Vito Luigi Colona, Maria Rosaria D’Apice, Paola Rogliani, Salvatore Zaffina, Francesca Leonardis, Andrea Campana, Massimiliano Raponi, Massimo Andreoni, Sandro Grelli, Giuseppe Novelli

**Affiliations:** Laboratory of Medical Genetics, Bambino Gesù Children’s Hospital, IRCCS, Rome, Italy; Department of Biology, Tor Vergata University of Rome, 00133 Rome, Italy; Medical Genetics Laboratory, Tor Vergata Hospital, Rome, Italy; Department of Biomedicine and Prevention, Tor Vergata University of Rome, 00133 Rome, Italy; Unit of Respiratory Medicine, Department of Experimental Medicine, University of Rome “Tor Vergata”, Rome, Italy; Occupational Medicine, Bambino Gesù Children’s Hospital, IRCCS, Rome, Italy; Intensive Care Unit, Tor Vergata University Hospital, Rome, Italy; Department of Pediatrics, IRCCS “Bambino Gesù” Children’s Hospital, Rome, Italy; Department of Systems Medicine, University of Rome Tor Vergata, Rome, Italy; Infectious Diseases Clinic, Policlinico Tor Vergata, Rome, Italy; Dept. of Experimental Medicine and Biochemical Sciences, University of Rome Tor Vergata, Italy; IRCCS Neuromed, Pozzilli (IS), Italy; Department of Pharmacology, School of Medicine, University of Nevada, Reno, NV 89557, USA

**Author notes:** Correspondence (G.N.).

## Abstract

**Background:** Coronaviruses (CoV) are a large family of viruses that are common in people and many animal species. Animal coronaviruses rarely infect humans with the exceptions of the Middle East Respiratory Syndrome (MERS-CoV), the Severe acute respiratory syndrome coronavirus (SARS-CoV), and now SARS-CoV-2, which is the cause of the ongoing pandemic of coronavirus disease 2019 (COVID-19). Many studies suggested that genetic variants in *ACE2* gene may influence the host susceptibility/resistance to SARS-CoV-2 virus according to the functional role of ACE2 in human pathophysiology. However, all these studies have been conducted *in silico* based on epidemiological and population data. We therefore investigated the occurrence of *ACE2* variants in a cohort of 99 Italian unrelated individuals clinically diagnosed with coronavirus disease 19 (COVID-19) to experimental demonstrate allelic association with disease severity.

**Methods:** By whole-exome sequencing we analysed 99 DNA samples of severely and extremely severely COVID-19 patients hospitalized at the University Hospital of Rome “Tor Vergata” and Bambino Gesù Hospital in Rome.

**Results:** We identified three different germline variants, one intronic (c.439+4G>A) and two missense (c.2158A>G, p.Asn720Asp; c.1888G>C, p.Asp630His), in 26 patients with a similar frequency between male and female and a not statistically different frequency, except for c.1888G>C, (p.Asp630His) with the ethnically matched populations (EUR).

**Conclusions:** Our results suggest that there is not any *ACE2* exonic allelic association with disease severity. It is possible that rare susceptibility alleles are located in the non-coding region of the gene able to control *ACE2* gene activity. It is therefore of interest, to explore the existence of *ACE2* susceptibility alleles to SARS-Co-V2 in these regulatory regions. In addition, we found no significant evidence that *ACE2* alleles is associated with disease severity/sex bias in the Italian population.

## Introduction

Coronaviruses (CoV) are a large family of viruses that are common in people and many animal species, including camels, cattle, cats, and bats. Animal coronaviruses rarely infect humans and then spread between people with the exceptions of the Middle East Respiratory Syndrome (MERS-CoV), the Severe acute respiratory syndrome coronavirus (SARS-CoV), and now SARS-CoV-2, which is the cause of the ongoing pandemic of coronavirus disease 2019 (COVID-19) (1; 2). SARS-CoV-2 utilizes an extensively glycosylated spike (S) protein that protrudes from the viral surface to bind to angiotensin-converting enzyme 2 (ACE2) to mediate host-cell entry (3). ACE2-binding affinity of the receptor-binding domain (RBD) in S1 subunit of S protein of SARS-CoV-2 is 10- to 20-fold higher than that of SARS-CoV, which may contribute to the higher infectivity and transmissibility of SARS-CoV-2 (4). Many studies suggested that genetic variants in *ACE2* gene may influence the host susceptibility/resistance to SARS-CoV-2 virus according to the functional role of ACE2 in human pathophysiology (2). However, all these studies have been conducted *in silico* based on epidemiological and population data. We therefore investigated the occurrence of *ACE2* variants in a cohort of 99 Italian SARS-CoV-2 -positive patients by direct exome sequencing.

## Results and Discussion

We analysed by exome sequencing 99 DNA samples of COVID-19 patients hospitalized at the University Hospital of Rome “Tor Vergata” in Rome and Bambino Gesù Hospital, Rome. All patients were clinically diagnosed as COVID-19 based on clinical suspicion and confirmed by viral RNA detection at oropharyngeal and nasopharyngeal swabs. All the selected patients were severely or extremely severely affected. Nine patients in the extremely severe group are passed away. The majority of the enrolled patients were males (61males, 38 females). Median age was 63 years (range: 2–92 years), 64 patients were under 65 years old. Ten patients were children (median age was 11.5 years) showing a severe form of the disease but none of them have Kawasaki-like syndrome (6). The analytical procedure received approval by the local ethics committee at University Hospital of Rome Tor Vergata (protocol no. 50/20).

We identified in *ACE2* gene, three different germline variants, one intronic (c.439+4G>A) and two missense (c.2158A>G, p.Asn720Asp; c.1888G>C, p.Asp630His), in 26 patients (14 females and 12 males). The frequency of the three variants identified are similar between male and female patients suggesting that there is not gender effects underlying the frequency distribution of *ACE2* variants. (**Table 1**).

**Table 1 -.**
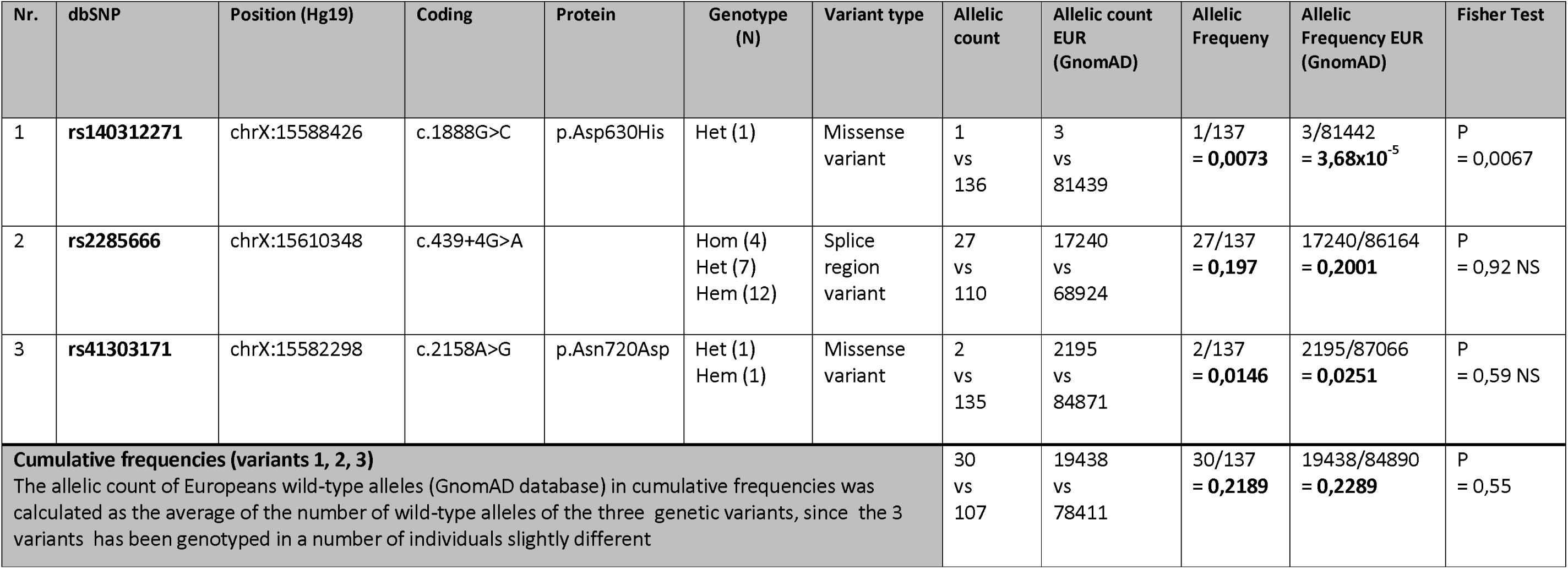
Comparison of allelic counts (variant vs WT alleles) between our Italian population of SARS-CoV2 Positive Patients and Europeans (GnomAD database)

| Nr. | dbSNP | Position (Hg19) | Coding | Protein | Genotype (N) | Variant type | Allelic count | Allelic count EUR (GnomAD) | Allelic Frequency | Allelic Frequency EUR (GnomAD) | Fisher Test |
| --- | --- | --- | --- | --- | --- | --- | --- | --- | --- | --- | --- |
| 1 | **rs140312271** | chrX:15588426 | c.1888G>C | p.Asp630His | Het (1) | Missense variant | 1<br>vs<br>136 | 3<br>vs<br>81439 | 1/137<br>= **0,0073** | 3/81442<br>= **3,68x10<sup>-5</sup>** | P<br>= 0,0067 |
| 2 | **rs2285666** | chrX:15610348 | c.439+4G>A |  | Hom (4)<br>Het (7)<br>Hem (12) | Splice region variant | 27<br>vs<br>110 | 17240<br>vs<br>68924 | 27/137<br>= **0,197** | 17240/86164<br>= **0,2001** | P<br>= 0,92 NS |
| 3 | **rs41303171** | chrX:15582298 | c.2158A>G | p.Asn720Asp | Het (1)<br>Hem (1) | Missense variant | 2<br>vs<br>135 | 2195<br>vs<br>84871 | 2/137<br>= **0,0146** | 2195/87066<br>= **0,0251** | P<br>= 0,59 NS |
| **Cumulative frequencies (variants 1, 2, 3)**<br>The allelic count of Europeans wild-type alleles (GnomAD database) in cumulative frequencies was calculated as the average of the number of wild-type alleles of the three genetic variants, since the 3 variants has been genotyped in a number of individuals slightly different |  |  |  |  |  |  | 30<br>vs<br>107 | 19438<br>vs<br>78411 | 30/137<br>= **0,2189** | 19438/84890<br>= **0,2289** | P<br>= 0,55 |

GnomAD database analysis revealed that the identified *ACE2* variants existed with a cumulative frequency of 0.2289 in ethnically matched populations (EUR). The frequency of the variants detected in our examined cohort (0.2119) was not statistically different (Table 1). A significant difference was detected only for the c.1888G>C (p.Asp630His) observed in a heterozygous female (p=0.0067) (Table 1). The allelic frequency of this variant in GnomAD for the EUR reference population is 0.000036 confirming that this is a very rare allele. To predict the potential impact of this variant on the protein we used different tools (PolyPhen2, Mutation Taster, SIFT, MetaLR_pred, and MetaSVM_pred.). The in-silico analyses gave conflicting computational verdict because 3 benign predictions vs. 2 pathogenic predictions. The sequence alignment of the ACE2 protein with its orthologous proteins shows that the wild type residue is not highly conserved in species implying an irrelevant functional or structural role of this residue in the ACE protein. Concerning the others two variants, the recurrent c.439+4A>G (rs2285666) intronic variant has been previously reported by Strafella et al. (5) and by Asselta et al.(4) in two different Italian cohort representative of the country’s population. The variant is located in the intron 3 in a splice site region of the gene. However, using Human Splicing Finder (HSF) the analysis showed no significant splicing alterations.

The missense variant c.2158A>G (p.Asn720Asp) was found in two patients, 1 female in heterozygous state and 1 male, with a frequency in line with the frequency reported for the European non-Finnish population in the GnomAD database. It is in the C-terminal domain that is not involved in the SARS-CoV-2 S protein interaction. The *in-silico* analyses to predict the potential impact of this variant on the protein gave benign computational verdict because 4 benign predictions vs 1 pathogenic prediction.

Numerous *in silico* data suggested that the ACE2 variants in structural part of the protein could have an impact on the pathogen binding dynamics or increase the quantitative expression of ACE2. All these studies were carried out on an epidemiological basis of population allele frequencies deposited in the various databases available. We systematically analyzed the *ACE2* coding-region variants in a representative cohort of Italian patients severely affected by COVID-19 in order to identify rare and causative predisposing alleles. Although we have identified a variant (p.Asp630His), very rare in European population, in a single patient affected by COVID-19, we do not believe that there is an enrichment of *ACE2* coding mutant alleles in the population of Italian patients affected by COVID-19. Our results confirm and extend that *ACE2* is a gene with low allelic frequencies of missense variants as expected on the basis of GnomAD population data. In fact, we provide evidence that the rate of amino acid changes in the binding region with SARS-Co-V2 and at the protein cleavage sites is very low. This suggests that these regions have been under evolutionary pressure, probably for the essential catalytic role of ACE2 as transmembrane carboxypeptidase. It is possible that rare susceptibility alleles are located in the non-coding region of the gene able to control *ACE2* gene activity. Mutant alleles in noncoding DNA can cause alteration in expression levels or disturbing the timing of the expression (1). These variations concern enhancers, promoters, insulators and silencers or regions that provide instructions for producing functional RNA molecules, such as transfer RNA, miRNAs or long non-coding RNA. It is therefore of interest, to explore the existence of *ACE2* susceptibility alleles to SARS-Co-V2 in these regulatory regions. Interestingly very recently Bunyavanich et al. (10) have showed age-dependent expression of ACE2 gene in nasal epithelium, highlighting how the different level of expression of ACE2 may be the reason for explaining why COVID-19 is less frequent in children (10). Several studies have shown that *ACE2* gene undergoes the action of at least four miRNAs: miR-200c, let-7b, miR-1246, and miR-125b (11-14). Genes coding for these miRNAs could reveal variations able to modulate the expression and therefore produce significant quantitative differences in the ACE2 protein.

## Data Availability

All data generated or analysed during this study are included in this published article.

## Acknowledgements

Thanks to Dr. Paolo Gravina, Dr. Laura Liberatoscioli and Valentina Lanari for their technical support. This work is part of the GEFACOVID Consortium.

## Web Resources

GnomAD, https://gnomad.broadinstitute.org/

PolyPhen2, http://genetics.bwh.harvard.edu/pph2/

Mutation Taster, http://www.mutationtaster.org/

SIFT, https://sift.bii.a-star.edu.sg/

